# ABO blood group type does not influence the level of SARS-CoV-2 antibody response in convalescent plasma donors

**DOI:** 10.1101/2021.10.19.21265195

**Authors:** Klemen Žiberna, Katerina Jazbec, Mojca Jež, Polonca Mali, Urška Rahne Potokar, Primož Rožman

## Abstract

The association of ABO blood group types with the COVID-19 disease has been confirmed by several studies, with the blood group A-type patients being more susceptible and prone to more severe clinical course of disease. Similarly, some authors explored the association of ABO-types and the levels of anti-SARS-CoV-2 antibodies in convalescents. The recent reports mostly support a theory that non-O blood group convalescents present with higher levels of anti-SARS-CoV-2 antibodies. Since these findings were based on small convalescent cohorts, we quantified the anti-SARS-CoV-2 antibodiy levels in four larger cohorts of total 3185 convalescent plasma donors with three commercial serological tests and one standard neutralizing antibody test. The majority of donors had undergone a mild form of disease and the median time of sampling was 66 days after the onset of COVID-19 symptoms. None of the antibody quantitation methods showed an association of the ABO blood group types with the level of anti-SARS-CoV-2 antibodies. The same result is evident in the group of vaccinated individuals (n=370) as well as in the groups stratified into three post-COVID-19 periods (0-60, 60-120, and 120-180 days). In conclusion we can state that the ABO blood group type does not influence the level of SARS-CoV-2 antibody response in COVID-19 convalescent plasma donors.

## Background

ABO histo-blood group antigens are expressed on blood cells, some other tissues, and in the body fluids. Already in the early studies of SARS-CoV-2 pandemic, the association of ABO blood group type with the SARS-CoV-2 infection has been noticed, where the blood group A was associated with an increased risk of disease compared to the blood group O that was associated with a decreased risk for severe COVID-19 illness^1, 2^. Although certain studies failed to confirm such association^3, 4^, several large cohort studies^5, 6^, reviews and meta-analyses^7-10^ confirmed that the individuals with blood group A are more susceptible and those with blood group O are less susceptible for COVID-19.

In the absence of specific anti-viral remedies, hyperimmune COVID-19 convalescent plasma (CCP) containing high titers of anti-SARS-CoV-2 antibodies (Abs) has been proposed as therapeutic modality early in the pandemic and it has been widely clinically used^11-14^. Because only CCP units with a high titer of anti-SARS-CoV-2 antibodies are recommended for treatment^15^, researchers have tried to define the factors affecting the quantity of anti-SARS-CoV-2 Abs in the convalescents. The correlation of antibody (Ab) levels in CCP donors with their ABO-type has been studied by several authors, who reported conflicting results mostly based on limited cohorts of COVID-19 convalescents. For instance, Li et al. reported that in 49 CCP convalescents, the levels of spike receptor binding domain (S-RBD)-specific and nucleocapsid (N)-specific IgG Abs had no significant correlation to their ABO-type^16^. Similarly, Körper et al. found no correlation between ABO-types and titers of neutralizing Abs in 144 CCP donors^17^, and Wendel et al. reported no such correlation in 78 CCP donors^18^. On the other hand, Madariaga et al. found in 103 CCP donors that AB-type donors had higher anti-RBD and anti-spike Ab levels compared to O-type donors^19^. De Freitas Dutra et al. reported in 268 donors and 162 patients, that O-type individuals had lower SARS-CoV-2 Ab titers than A and AB individuals^20^. Bloch et al. reported that in 202 CCP donors there were no significant differences in anti-spike IgA or anti-spike IgG Ab titers by ABO blood group, but that significantly more B-type individuals presented with a high neutralization Ab titer (≥1:160) compared to O-type and A-type donors^21^. In a recent paper Hayes et al. reported that in 232 O-type convalescents they found significantly lower levels of SARS-CoV-2 IgG Abs compared to A-type individuals^22^.

Based on these ambiguities we decided to clarify this question by exploring the association of ABO-type with the concentration of anti-SARS-CoV-2 antibodies and neutralizing antibodies in a much larger cohort of 3185 CCP donors.

## Methods

### Convalescent plasma donors

The first donations from 3185 CCP donors with a history of polymerase chain reaction (PCR) confirmed SARS-CoV-2 infection in nasopharyngeal swab were included and collected between June 2020 and August 2021. Standard eligibility criteria for volunteer blood donors were used. Out of these, 90% participants reported having mild symptoms and 10% reported moderate to severe symptoms with 6 participants requiring hospital treatment. The median (IQR) duration between the start of COVID-19 symptoms and CCP donation was 66 (45-114) days.

The ABO blood group distribution in the 3185 CPP donors was 40.5% A, 37.9% O, 14.9% B, and 6.8% AB, which matches our 2017 data for general blood donor population^23^. Since the vaccination has been implemented after January 2021, 370 convalescent donors have also received the vaccination before their first CPP collection. Basic donor information is presented in Table 1.

**Table 1.** Basic demographic factors, results of serology tests, and vaccination status in COVID-19 convalescent plasma donors with different ABO blood group types.

|  | **Group O**<br>**(N=1206)**<br>**37.9%** | **Group A**<br>**(N=1289)**<br>**40.5%** | **Group B**<br>**(N=475)**<br>**14.9%** | **Group AB**<br>**(N=215)**<br>**6.8%** | **p-value** |
| --- | --- | --- | --- | --- | --- |
| Gender - female | 423 (35.1%)<br>(N=1206) | 447 (34.7%)<br>(N=1289) | 169 (35.6%)<br>(N=475) | 71 (33.0%)<br>(N=215) | 0.925 |
| Age (years) | 41.94±0.32<br>(N=1206) | 41.33±0.30<br>(N=1289) | 42.29±0.52<br>(N=475) | 42.15±0.73<br>(N=215) | 0.300 |
| Body weight (kg) | 83.97±0.49<br>(N=1050) | 84.41±0.49<br>(N=1124) | 83.99±0.8<br>(N=420) | 84.39±1.22<br>(N=182) | 0.923 |
| Days after start of COVID-19 symptoms | 65<br>[45-110]<br>(N=1116) | 66<br>[45-115]<br>(N=1177) | 66<br>[46-120]<br>(N=438) | 73<br>[43-115]<br>(N=198) | 0.838 |
| Wantai semi-quantitative SARS-CoV-2 Ab test (index S/C) | 15.82<br>[8.29-18.95]<br>(N=158) | 17.71<br>[8.02-19.19]<br>(N=178) | 17.84<br>[11.09-19.47]<br>(N=67) | 18.59<br>[9.61-19.71]<br>(N=31) | 0.224 |
| Abbott semi-quantitative SARS-CoV-2 Ab test (index S/C) | 3.81<br>[1.90-5.71]<br>(N=380) | 3.63<br>[1.84-5.89]<br>(N=401) | 3.68<br>[1.92-5.87]<br>(N=161) | 3.97<br>[1.3-5.72]<br>(N=77) | 0.998 |
| Abbott quantitative SARS-CoV-2 Ab test (AU/ml) | 451<br>[194-1694]<br>(N=994) | 558<br>[212-2160]<br>(N=1047) | 510<br>[221-2751]<br>(N=355) | 472<br>[169-1953]<br>(N=164) | 0.094 |
| Neutralization test (titer) | 80<br>[40-320]<br>(N=417) | 80<br>[20-320]<br>(N=558) | 80<br>[20-320]<br>(N=223) | 80<br>[20-320]<br>(N=104) | 0.414 |
| Nr. of units with high neutralization test values (>160) | 159 (38.1%)<br>(N=417) | 239 (42.8%)<br>(N=558) | 87 (39.0%)<br>(N=223) | 38 (36.5%)<br>(N=104) | 0.382 |
| Vaccination | 126 (10.4%) | 157 (12.2%) | 64 (13.5%) | 23 (10.7%) | 0.287 |

**Legend:** Data are presented as a mean and a standard error of mean (SEM) for normally distributed data, a median and an interquartile range (IQR) for non-parametric data, and as a count and a percentage for binary data. The statistical comparison between multiple groups was performed using one-way ANOVA test for normally distributed data, Kruskal-Wallis ANOVA test for non-parametric data, and Pearson's chi-squared test for binomial data.
\*S/C, sample/cut-off; AU, arbitrary units

### Serology testing

Two semi-quantitative and one quantitative serological tests were used to detect anti-SARS-CoV-2 antibodies: i) Wantai SARS-CoV-2 Ab ELISA, an enzyme-linked immunosorbent assay for qualitative detection of total IgG and IgM antibodies to the RBD of SARS-CoV-2 spike protein that was performed on 434 samples; ii) Abbott SARS-CoV-2 IgG assay, a chemiluminescent microparticle assay (CMIA) for the qualitative detection of IgG Abs to the nucleocapsid protein that was performed on 1019 samples; and iii) Abbott SARS-CoV-2 IgG II Quant, second generation CMIA for quantitative determination of IgG Abs to the RBD of the S1 subunit of the SARS-CoV-2 spike protein, including the neutralizing Abs that was performed on 2560 samples. All tests were performed according to the manufacturers’ instructions.

For the neutralizing antibody testing, a standard live SARS-CoV-2 microneutralization assay was used^24^. The assay read out was the cytopathic effect, where assay cut-off titer was <1:20. Neutralization test was performed on 1302 samples by the Institute of Microbiology and Immunology, Faculty of Medicine, University of Ljubljana.

## Results and Discussion

The antibody analysis according to different ABO blood group types is presented in **Table 1**. There were no statistically significant differences between the SARS-CoV-2 antibody levels in the different ABO-types regardless of which serological method was used.

Although the quantitative Abbott SARS-CoV-2 IgG II Quant test resulted in slightly higher titers in the A-type group (558 AU/mL), the differences compared to other blood group types are not significant. The same dynamic is seen in the neutralization test titer values: there are no differences between the donors of different ABO blood group types. The proportion of samples with high neutralization test values (titer >160) reveals no ABO-type related differences either.

If the cohort is divided into vaccinated (n=370) and unvaccinated convalescents (n=2815), the result is similar and no statistically significant differences are evident. However, average titers of IgG and neutralizing antibodies in the vaccinated groups were as expected significantly higher (1:1,280 [1:640-11,280] vs. 1:80 [1:20-1:160], p<0.001) in the neutralization tests, and 1:12,200 vs. 1:380 AU/mL in the Abbott quantitative test (data not shown).

As there is a possibility that analysis of CCP donations in different period post infection would lead to different results, we analyzed antibody responses in unvaccinated CCP donors grouped by different donation periods after the onset of COVID-19 symptoms, which led to the same conclusion: there are no statistically significant differences between donors of different ABO blood groups (see **Table 2**).

**Table 2.** Antibody response in unvaccinated CCP donors of different ABO-types according to the post-infection period as measured by quantitative and neutralization tests.

|  | O | A | B | AB | p-value |
| --- | --- | --- | --- | --- | --- |
| **Abbott quantitative SARS-CoV-2 Ab test (AU/mL)** |  |  |  |  |  |
| 0 - 60 days after start of COVID-19 symptoms | 418.5<br>[187.0-968.8]<br>(N=345) | 446.5<br>[189.4-1038.57]<br>(N=354) | 405.95<br>[204.7-848.15]<br>(N=118) | 532.3<br>[166.02-1587.73]<br>(N=44) | 0.995 |
| 60 - 120 days after start of COVID-19 symptoms | 340.5<br>[165.8-825.77]<br>(N=318) | 402.45<br>[192.6-908.15]<br>(N=334) | 395.0<br>[170.25-963.65]<br>(N=103) | 357.15<br>[192.47-782.15]<br>(N=58) | 0.605 |
| 120 - 180 days after start of COVID-19 symptoms | 279.0<br>[135.07-690.0]<br>(N=110) | 421.5<br>[152.9-1161.3]<br>(N=101) | 325.25<br>[138.12-694.1]<br>(N=38) | 161.85<br>[107.42-405.35]<br>(N=22) | 0.145 |
| **Neutralization test (titer)** |  |  |  |  |  |
| 0 - 60 days after onset of COVID-19 symptoms | 40.0<br>[20.0-160.0]<br>(N=192) | 40.0<br>[20.0-160.0]<br>(N=215) | 40.0<br>[0.0-80.0]<br>(N=97) | 40.0<br>[5.0-80.0]<br>(N=50) | 0.593 |
| 60 - 120 days after onset of COVID-19 symptoms | 60.0<br>[40.0-80.0]<br>(N=96) | 40.0<br>[20.0-80.0]<br>(N=128) | 80.0<br>[20.0-160.0]<br>(N=52) | 40.0<br>[5.0-80.0]<br>(N=22) | 0.637 |
| 120 - 180 days after onset of COVID-19 symptoms | 80.0<br>[20.0-80.0]<br>(N=28) | 80.0<br>[20.0-160.0]<br>(N=40) | 40.0<br>[20.0-80.0]<br>(N=13) | 30.0<br>[15.0-50.0]<br>(N=4) | 0.586 |

**Legend:** Data are presented as a median and an interquartile range (IQR) and the statistical comparison between groups was performed using Kruskal-Wallis ANOVA test. AU, arbitrary units.

In conclusion, by using a large cohort of convalescents and several antibody quantification methods, we found no evidence to confirm some earlier reports that ABO blood group type impacts the quantity of anti-SARS-CoV-2 antibodies in COVID-19 convalescent donors of hyperimmune plasma.

## Data Availability

All data produced in the present study are available upon reasonable request to the authors

## Declarations

### Ethics

The study was approved by the national Medical Ethics Committee of Republic of Slovenia (0120-241/2020-8, from 18.6.2020) and conducted from June 2020 to August 2021.

## Authorship

### Contribution

KŽ collected, verified, and analyzed the data, managed the software, and wrote the draft manuscript. MJK and KJ collected data, investigated methodology, wrote the draft, and revised the manuscript. PM acquired the funding, supervised, and administered the project, and critically reviewed the manuscript. URP analyzed the samples and wrote the manuscript. PR acquired the funding, conceptualized, designed, and supervised the study, and wrote the manuscript.

All authors read and approved the final manuscript.

## Acknowledgements

Authors would like to thank Maja Černilec and Marjana Šprohar for help with collecting and distributing the samples; Petra Jovanovič and Sonja Vuletić for help with analyzing the samples, to all personnel at Blood Transfusion Centre of Slovenia for their contribution to the collection of CCP donations, and to Stephen Minger for critical reading of the letter.

